# A Framework for Evaluating the Efficacy of Foundation Embedding Models in Healthcare

**DOI:** 10.1101/2024.04.17.24305983

**Authors:** Sonnet Xu, Haiwen Gui, Veronica Rotemberg, Tongzhou Wang, Yiqun T. Chen, Roxana Daneshjou

## Abstract

Recent interest has surged in building large-scale foundation models for medical applications. In this paper, we propose a general framework for evaluating the efficacy of these foundation models in medicine, suggesting that they should be assessed across three dimensions: general performance, bias/fairness, and the influence of confounders. Utilizing Google’s recently released dermatology embedding model and lesion diagnostics as examples, we demonstrate that: 1) dermatology foundation models surpass state-of-the-art classification accuracy; 2) general-purpose CLIP models encode features informative for medical applications and should be more broadly considered as a baseline; 3) skin tone is a key differentiator for performance, and the potential bias associated with it needs to be quantified, monitored, and communicated; and 4) image quality significantly impacts model performance, necessitating that evaluation results across different datasets control for this variable. Our findings provide a nuanced view of the utility and limitations of large-scale foundation models for medical AI.

## 1. Introduction

There is a global need for accessible medical care. A shortage of experts, especially in under-resourced countries, presents an opportunity to develop automated medical tools to assist this process through the use of artificial intelligence (AI). Deep learning approaches have demonstrated the ability to perform accurate generalization on highly variable medical classification tasks within fields such as dermatology (Esteva et al., 2017; Han et al., 2020b; Tschandl et al., 2018). However, current methods such as the Convolutional Neural Network (CNN) are computationally expensive to train and rely on large amounts of domain-specific data. Medical image and data scarcity due to factors ranging from quality control, and patient privacy, to the expensive cost of expert annotations, present a prohibitively large barrier to creating the large datasets necessary to train field-specific models. This makes the creation of domain-specific tools difficult.

One proposed solution to this is the use of foundation models - large-scale models trained on large corpora of data that can then be fine-tuned on downstream tasks (Gui et al., 2024). While the initial training of foundation models is data-hungry and computationally expensive, by learning general features from a wide range of data sources, they offer the potential to reduce the data, computing, and technical expertise necessary for building models from scratch for every task.

However, the use of these foundation models in other domains such as language modeling has revealed that foundation models can hold intrinsic biases against certain demographic groups, in regard to aspects such as race, which limits their applicability. The lack of transparency about input data sources for the training of such models also raises concerns surrounding data diversity and representation. Because of this, their performance in novel tasks can be unpredictable and variable.

This paper pilots a framework for evaluating the efficacy of foundation models in medicine. We suggest that these models should be evaluated through three dimensions: general performance, bias/fairness, and influence of confounders. Through the use of Google’s recently released dermatology embedding model (Steiner, 2024), we demonstrate this new framework by specifically looking at general performance (Sections 3.2.1 and 3.2.2), performance across skin tones (Section 3.2.3), and the impact of photo quality as a potential confounder (Section 3.2.4). First, we evaluate the effect of skin tone on internal representation, by generating over 10,000 embeddings for the International Skin Imaging Collaboration (ISIC) and Diverse Dermatology Images (DDI) datasets and comparing how similar their representations are across Fitzpatrick categories and skin conditions. Second, we evaluate performance by building linear classification models with the generated embeddings to assess the utility of these embeddings in downstream diagnostic tasks, comparing the performance to different zero-shot models. Third, we assess the impact of differing image quality on the foundation model as a potential confounder.

### Generalizable Insights about Machine Learning in the Context of Healthcare

Our study proposes a new framework for evaluating the applicability of emerging foundation models to clinical tasks to help medical users. Our findings highlight the importance of evaluating robustness across three areas, specifically downstream performance, potential for bias, and possible confounding effects.

Our work presents the following generalizable insights for machine learning and healthcare: 1) Skin color is a crucial internal differentiator for machine learning models, emphasizing the need to evaluate and address this potential bias; 2) General-purpose CLIP models encode substantial understanding of images relevant to medical tasks such as lesion diagnostics in dermatology. This suggests that these models serve as a strong baseline and building block for specialized medical AI efforts.

## 2. Related Work

### 2.1. Deep Learning in Healthcare

In the past decade, there has been an increase in applications of deep learning techniques, including computer vision, natural language processing, and reinforcement learning, to healthcare. In particular, the field has seen tremendous progress in applying computer vision models to segmentation, detection, and classification of images in the medical setting (Esteva et al., 2019). One of the most popular and successful algorithms in computer vision is the convolutional neural networks (CNN), which has been found to achieve physician-level accuracy at a broad variety of diagnostic tasks, including identifying diabetic retinopathy from images of the eye (Gulshan et al., 2016), detecting breast lesions in mammograms (Kooi et al., 2017), and analyzing spinal MRI images (Jamaludin et al., 2017).

In dermatology, multiple deep learning approaches have been developed, including ModelDerm (Han et al., 2020b), DeepDerm (Esteva et al., 2017), and HAM10000 (Tschandl et al., 2020). These models have all previously demonstrated state-of-the-art (SOTA) performance in diagnosing lesions from images of the skin, and have similarly been found comparable to dermatologists’ performance (Haenssle et al., 2020; Esteva et al., 2017). Yet, despite the impressive performance by these SOTA algorithms, several of these algorithms have been shown to be poorly generalizable, resulting in variable performances when tested outside of the original experimental conditions (Du-Harpur et al., 2020).

### 2.2. Foundation Models in Healthcare

To help mitigate some of the limitations of traditional deep learning models, foundation models have become more popular (Bommasani et al., 2022). These large-scale, computationally expensive and data-hungry models serve as the foundation for additional models to be built upon it. Using transfer learning, the base foundation model can be adjusted for specific tasks if needed. Unlike deep learning where large amounts of task-specific data must be available for the model to learn from, foundation models create broader general-purpose features that can be used in multiple scenarios.

Google has recently released Derm Foundation, an foundational embedding model that is derived from deep learning applications to dermatology images (Steiner, 2024). Derm Foundation is based on a BiT ResNet-101x3 that was trained in 2 stages. The first pretraining stage used contrastive learning to train on a large number of image-text pairs; the second stage fine-tuned this base model for diagnostic classification using clinical datasets. The team found that models built on top of the Derm Foundation embeddings for dermatology-related tasks achieved significantly higher quality than previous models, demonstrating that Derm Foundation can serve as a useful starting point to accelerate dermatology-related modeling tasks.

### 2.3. Biases in AI Models in Dermatology

Many AI models in dermatology suffer from lack of skin tone representation in training and testing datasets, resulting in biased performance on different Fitzpatrick skin tones. Some studies only validate models on a single race from a specific region (Han et al., 2020a). Previous work showed SOTA models performed significantly worse on images from patients with Fitzpatrick V/VI (Daneshjou et al., 2022, 2021). One case study showed that only 17% of FST VI images were correctly diagnosed through models that were predominately trained on light skin types (Kamulegeya et al., 2023).

## 3. Methods

### 3.1. Datasets

We evaluate the embeddings of dermatology foundation models on two different datasets: the Diverse Dermatology Imgaes Dataset (DDI), and the International Skin Imaging Collaboration (ISIC) 2018 dataset. DDI is a benchmark dataset with 656 images of skin disease across skin tones with labels confirmed via biopsy and histopathology (Daneshjou et al., 2022). Each image includes the biopsy-proven diagnosis in addition to a broad categorization of lesions into malignant and benign. Each image also contains information about the Fitzpatrick skin type (FST), a scale that is frequently used in dermatology to quantify skin tones (Fitzpatrick, 1988). The dataset was designed to allow matching patient characteristics for direct comparison between patients classified as FST V–VI (dark skin tones) and patients with FST I–II (light skin tones). The FST I–II, III-IV, and V-VI subgroups in DDI consist of 208 images (159 benign and 49 malignant), 241 images (167 benign and 74 malignant), and 207 images (159 benign and 48 malignant), respectively. DDI also included an image quality score, which was the mean from independent assessments by three board-certified dermatologists. The initial image quality scores range from 0 (highest quality) to 4 (lowest quality), but images with a score of 4 were removed from the final dataset.

The ISIC 2018 dataset (Tschandl et al., 2018; Codella et al., 2019) consists of 10,015 dermatoscopic images of pigmented lesions collected from Austria and Australia. The dataset includes seven diagnostic categories: 1) 327 cases of actinic keratoses and intraepithelial carcinoma (akiec), 2) 514 cases of basal cell carcinoma (bcc), 3) 1,099 cases of benign keratosis (bkl), 4) 115 cases of dermatofibroma (df), 5) 6,705 cases of melanocytic nevi (nv), 6) 1,113 cases of melanoma (mel), and 7) 142 cases of vascular lesions (vasc). More than 50% of these lesions were confirmed by pathology, and the ground truth for the remaining cases was determined via follow-up, expert consensus, or confirmation by *in vivo* confocal microscopy (Tschandl et al., 2018). In addition to one-versus-others classification for the seven categories of diagnosis, we also grouped the diagnostic categories into malignant (akiec, bcc, mel) and benign (bkl, df, nv, and vasc) lesions.

Based on personal correspondence on 3/27/2024 with Yuan Liu, an author of the Google Derm Foundation embedding model, Derm Foundation was not trained on either DDI or ISIC 2018 datasets.

### 3.2. Evaluating Foundation Model Embeddings

#### 3.2.1. General predictive performance of foundation models

We primarily evaluated the performance of Derm Foundation, a fine-tuned BiT-M model with an embedding size of 6,144. To analyze the diagnostic performance of the foundation models, we first generated corresponding image embeddings for both DDI and ISIC and evaluated the performance of predicting the malignant versus benign lesion outcome using a simple logistic regression classifier on top of the embeddings (with default penalty parameter as implemented in `scikit-learn` (Pedregosa et al., 2011). We report the five-fold cross-validated AUC, accuracy, precision, recall, specificity, and F1-score using the implementation in `scikit-learn`. For the classification task on DDI, we conducted additional analysis on disjoint training and testing sets (i.e., no overlapping patients) to reduce data leakage.

#### 3.2.2. Comparing Representation Across Different Models

We compared the performance of Derm Foundation embeddings with embeddings derived from one additional dermatology model MONET (Kim et al., 2024), a fine-tuned CLIP model with an embedding size of 768, as well as 78 other non-domain-specific vision models, following the work of Huh et al. (2024). These models include ViT (Dosovitskiy et al., 2020), ResNet-50 (He et al., 2016), and ResNeT-18 (He et al., 2016) (the full list of models considered can be found in Appendix A). We computed the five-fold cross-validated metrics for the same set of predictive tasks (malignant or benign) for all embeddings considered.

To measure the similarities between the model embeddings, we used a nearest-neighbor metric in Huh et al. (2024) that computes the average overlap between *k*-nearest-neighbors for each embedding, which has been shown to capture model similarities. To account for the drastically different dataset sizes, we set *k* to 6 for DDI and 98 for ISIC.

#### 3.2.3. Examining the potential bias for different skin tone subgroups

Given the demonstrated disparity in many existing models’ performance on downstream diagnostic tasks across subgroups with different Fitzpatrick Skin Tones, we explored how the predictive performance and embedding geometries vary across three different FST subgroups (I–II, III–IV, and V–VI; from lightest to darkest skin tones). The DDI dataset was designed to allow a demographic parity fairness assessment by allowing direct comparisons between FST I–II and V–VI, which were matched.

For geometrical investigations, we computed cosine similarities between benign and malignant samples, a proxy for diagnostic values, across different subgroups. In addition, we also examined the performance of the *𝓁* _1_-regularized logistic regression classifier trained on different FST subgroups to understand the generalizability of the learned embeddings.

#### 3.2.4. Exploring confounding factors, including image quality

We also assessed the effects of image quality on the predictive performance of the embeddings. Using DDI’s image quality score, we defined high-quality and low-quality images to be those with quality scores *<* 1 and *>* 1.5, respectively. We used *𝓁* _2_-regularized logistic regressions to assess the test set performance by training on high or low-quality images only.

## 4. Results

### 4.1. Classification accuracy using foundation model embeddings

For DDI, we found that using Derm Foundation embeddings with a simple logistic classifier produced competitive results, with an AUROC of 0.76, a precision of 0.72, and a recall of 0.58 (see Table 1). For context, Daneshjou et al. (2022) reported AUROCs ranging from 0.56 to 0.67 for dermatology models on the same task. For ISIC 2018, using Derm Foundation embeddings yields an AUROC of 0.78, a recall of 0.61, and a specificity of 0.94; these numbers are competitive with previously reported performance. For example, Tschandl et al. (2020) reported a recall of 0.59 and a specificity of 0.93. Our results confirm that Derm Foundation embeddings contain informative features for diagnostic tasks.

**Table 1:** Predictive performance of using an *𝓁* _2_ logistic regression on Derm Foundation embeddings to predict malignant versus benign lesions on DDI (first row) and ISIC (second row), as well as performance on predicting specific diagnoses (one-versus-rest) on ISIC (third to last rows).

| Dataset | Class | Specificity | Precision | Recall | F1-score | AUROC | Accuracy |
| --- | --- | --- | --- | --- | --- | --- | --- |
| DDI | Malignancy | 0.94 | 0.82 | 0.58 | 0.68 | 0.76 | 0.83 |
| ISIC 2018 | Malignancy | 0.94 | 0.71 | 0.61 | 0.66 | 0.78 | 0.88 |
| ISIC 2018 | akiec | 0.99 | 0.65 | 0.53 | 0.58 | 0.76 | 0.53 |
| ISIC 2018 | bcc | 0.99 | 0.75 | 0.67 | 0.70 | 0.83 | 0.67 |
| ISIC 2018 | bkl | 0.96 | 0.70 | 0.67 | 0.69 | 0.82 | 0.67 |
| ISIC 2018 | df | 1.00 | 0.94 | 0.65 | 0.77 | 0.83 | 0.65 |
| ISIC 2018 | mel | 0.96 | 0.66 | 0.54 | 0.59 | 0.75 | 0.54 |
| ISIC 2018 | nv | 0.80 | 0.90 | 0.96 | 0.93 | 0.88 | 0.96 |
| ISIC 2018 | vasc | 1.00 | 0.84 | 0.91 | 0.87 | 0.96 | 0.91 |

### 4.2. Performance of Different Vision Foundation Models

In Figure 1, we display the average AUC across five-fold cross-validation for all vision models considered in Section 3.2.2 in a two-dimensional UMAP plot for both DDI (left panel of Figure 1) and ISIC (right panel of Figure 1). We note that in both cases, dermatology foundation models (both Derm Foundation and MONET) attain the highest AUC compared to models trained on natural images and texts only. However, the gap between these dermatology foundation models (depicted as stars in Figure 1) and general-purpose CLIP foundation models (depicted as crosses in Figure 1) is quite small (*<* 2% AUC). Moreover, we note that models trained with *only* natural images or synthetic images perform much worse on the downstream prediction task. Our observations indicate that image-text paired training, even using only general-purpose images, might provide generalization for domain-specific downstream tasks in dermatology.

**Figure 1:**
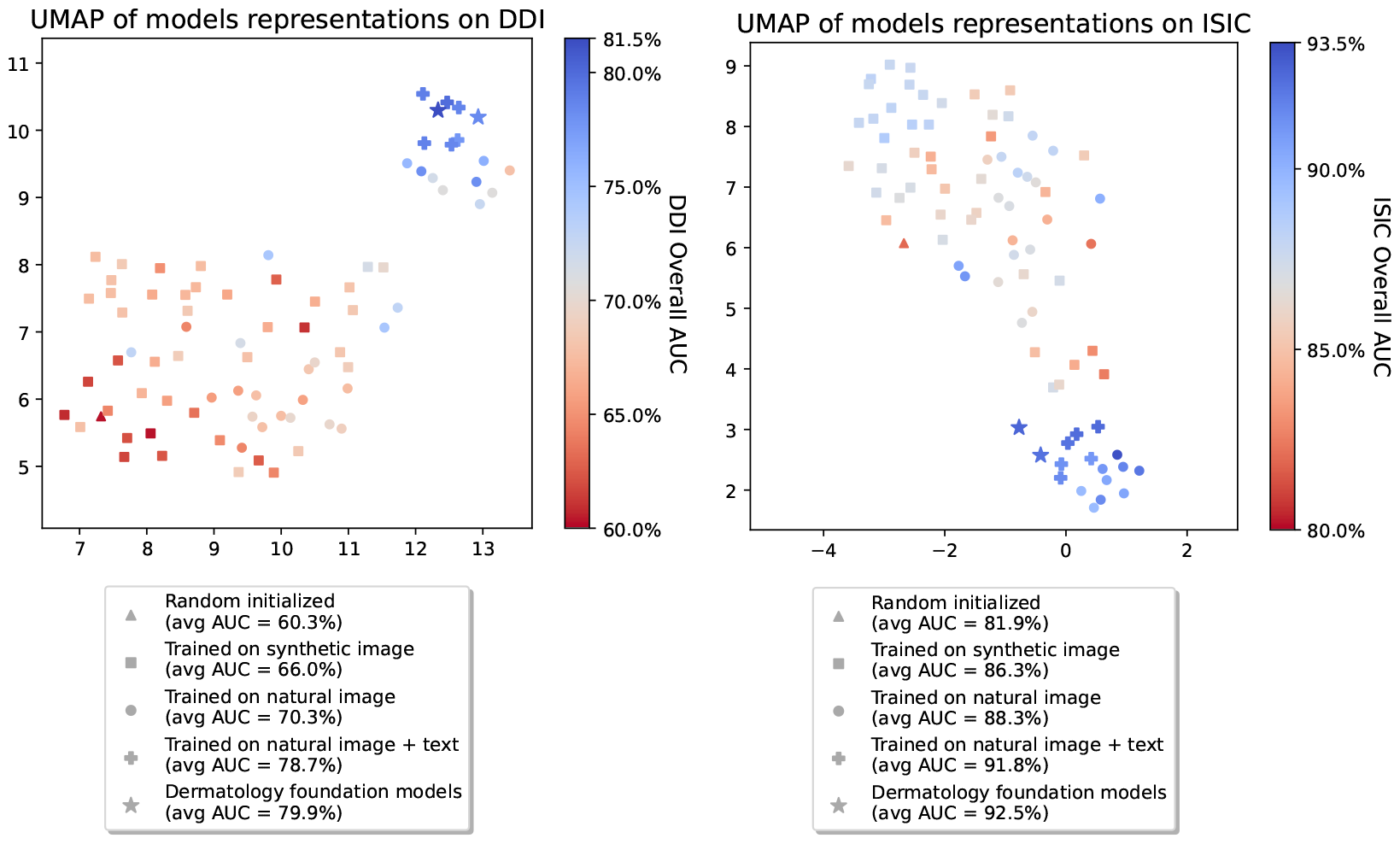
UMAP visualization of vision model embeddings on DDI (left) and ISIC (right). Each point corresponds to a different model (details listed in Appendix A) and is displayed in different shapes according to classes of models and colored by the mean AUC across five-fold cross-validation.

Finally, we observe that better-performing models are generally more clustered together in the 2D visualization than other models, akin to the previously documented “Anna Karenina Principle”, in which less generalizable models (i.e., “unhappy families”) vary more in the corresponding representation than more generalizable models (i.e., “happy families”) (Ainsworth et al., 2022; Bansal et al., 2021). This aligns with the conclusions in Huh et al. (2024), which tested with natural images rather than domain-specific medical ones.

### 4.3. Differential Predictive Performance of Embedding Models in Different Skin Tone Groups

In Table 2, we evaluated the predictive performance of foundation model embeddings on subgroups of different skin tones, as well as the generalizability and potential bias of these embeddings by testing whether a model learned on one subgroup could be used to achieve high predictive performance for another group.

**Table 2:** Predictive performance of using an *𝓁* _2_ logistic regression on Derm Foundation embeddings to predict malignant versus benign lesions, stratified by training subset and testing subset (I/II, III/IV, V/VI denote the subsets of DDI images from patients with Fitzpatrick scale I-II, III-IV, and V-VI, respectively). a: For the entries where train and test are the same subsets, metric averages over five-fold cross-validation are reported as a reference.

| Train | Test | Specificity | Precision | Recall | F1 | AUC | Accuracy |
| --- | --- | --- | --- | --- | --- | --- | --- |
| I/II<sup>a</sup> | I/II | $0.88 \pm 0.05$ | $0.56 \pm 0.12$ | $0.49 \pm 0.17$ | $0.69 \pm 0.08$ | $0.78 \pm 0.07$ | $0.79 \pm 0.06$ |
| III/IV | I/II | 0.72 | 0.41 | 0.63 | 0.64 | 0.76 | 0.70 |
| V/VI | I/II | 0.93 | 0.54 | 0.27 | 0.61 | 0.68 | 0.77 |
| III/IV<sup>a</sup> | III/IV | $0.89 \pm 0.04$ | $0.73 \pm 0.06$ | $0.62 \pm 0.11$ | $0.76 \pm 0.03$ | $0.83 \pm 0.05$ | $0.81 \pm 0.02$ |
| I/II | III/IV | 0.84 | 0.65 | 0.66 | 0.75 | 0.82 | 0.79 |
| V/VI | III/IV | 0.94 | 0.67 | 0.27 | 0.61 | 0.69 | 0.73 |
| V/VI<sup>a</sup> | V/VI | $0.93 \pm 0.02$ | $0.72 \pm 0.06$ | $0.56 \pm 0.09$ | $0.76 \pm 0.04$ | $0.82 \pm 0.04$ | $0.85 \pm 0.02$ |
| I/II | V/VI | 0.73 | 0.36 | 0.50 | 0.60 | 0.68 | 0.68 |
| III/IV | V/VI | 0.82 | 0.37 | 0.35 | 0.59 | 0.68 | 0.71 |

First of all, when comparing the predictive performance within subgroups on a train/test split only, the predictive performances are comparable, with higher accuracy and performance observed in both the III/IV and V/VI subgroups.

The trend of training and testing on different FST subgroups is more nuanced. We note that when testing on patients with the lightest skin tone (the I/II subgroup), we see overall comparable performance but an increase in recall and a drop in precision (from 0.56 to 0.41) when the training sample consists of a similar but different subgroup (III/IV). However, there is a significant change in recall (from 0.49 to 0.27) when training samples are from the more distinctly different dark skin groups. Interestingly, a similar observation holds when the test samples are from III/IV only as well: we see a noticeable drop in recall when training only on V/VI. On the other hand, if V/VI (the darkest subgroup) is of interest, training on I/II and III/IV led to noticeable drops in both precision and recall.

To further understand the generalization experiment results, we display average cosine similarities between Derm Foundation embeddings across different diagnostic classes and Fitzpatrick Skin Tones in Table 3. We note several overall trends: 1) the Fitzpatrick Skin Tone (FST) dominates the cosine similarity calculation, i.e., even for samples with the same diagnostic classes, different skin tones reduce the average cosine similarity to below 0.4; 2) overall, given samples from the same FST class, benign samples are most similar to each other, followed by malignant samples, and then the overall samples.

**Table 3:** Average cosine similarities between Derm Foundation embeddings across different diagnostic classes and Fitzpatrick Skin Tones.

| FST Categories | Classes | Average Cos Sim |
| --- | --- | --- |
| I/II & I/II | benign & benign | 0.55 |
| I/II & I/II | benign & malignant | 0.50 |
| I/II & I/II | malignant & malignant | 0.53 |
| I/II & V/VI | benign & benign | 0.37 |
| I/II & V/VI | malignant & malignant | 0.32 |
| V/VI & V/VI | benign & benign | 0.48 |
| V/VI & V/VI | benign & malignant | 0.41 |
| V/VI & V/VI | malignant & malignant | 0.43 |

### 4.4. Evaluating Image Quality as a Potential Confounder for Performance

In Table 4, we reported the performance of an *𝓁* _2_-regularized logistic regression model using Derm Foundation embeddings on different combinations of train and test subsets consisting of high- and low-quality images, as image quality could act as a coufounder when evaluating downstream prediction accuracy. We note that overall, if the test set consists only of high-quality images, then training on either high or low-quality images would lead to similar predictive performance (in fact, marginally higher if trained on low-quality images). On the other hand, the predictive performance is much more sensitive to the training set composition if we want to draw inferences on the low-quality images. In particular, training on high-quality images improves the test performance on low-quality images by a large margin (around 0.2 improvement across precision, recall, F1, and AUROC) compared to training and testing on low-quality images. That said, overall, the results on the DDI dataset are quite robust to the quality of the training images, likely due to the removal of the lowest quality tier during dataset construction.

**Table 4:** Predictive performance of using an *𝓁* _2_ logistic regression on Derm Foundation embeddings to predict malignant versus benign lesions, stratified by training and testing subsets (High and Low denote the subsets of DDI images of high and low quality, as assessed by dermatologists, respectively).

| Train | Test | Specificity | Precision | Recall | F1-score | AUROC | Accuracy |
| --- | --- | --- | --- | --- | --- | --- | --- |
| High | High | 0.90 | 0.50 | 0.33 | 0.40 | 0.62 | 0.78 |
| High | Low | 0.91 | 0.58 | 0.50 | 0.54 | 0.71 | 0.83 |
| Low | High | 0.86 | 0.63 | 0.54 | 0.58 | 0.70 | 0.76 |
| Low | Low | 0.85 | 0.40 | 0.25 | 0.31 | 0.55 | 0.68 |

## 5. Discussion

### 5.1. Improvement over SOTA

Compared to previous SOTA algorithms, dermatology foundation models such as Derm Foundation and MONET achieve higher classification accuracy. In particular, the improved performance on diverse datasets such as DDI, which often poses difficulty for previous models, suggests that foundation models may hold promise in medical applications.

### 5.2. Domain-specific versus General Foundation Models

We also find that the improvement over the previous state-of-the-art (SOTA) persists for general-purpose CLIP models (see Figure 1). This indicates that image-text paired training is likely the key ingredient for success. Consequently, general-purpose CLIP models should be more broadly considered as a baseline for medical tasks, such as lesion diagnostics in dermatology. Domain-specific models should be compared to and developed based on these performant general-purpose models, which are often open-source and come with extensive resources on deployment.

### 5.3. Skin-tone is a key diffrentiator

Interestingly, we found that for the embeddings of Derm Foundation, there were higher cosine similarities between benign and malignant skin conditions of the same FST category, than between FST categories with the same classification label. Although this did not seem to adversely affect accuracy, this suggests that machine learning models cannot perform race-agnostic diagnoses and implicitly factor skin tone into internal evaluations.

### 5.4. Confounding Effects of Image Quality

Unsurprisingly, image quality significantly impacts model performance — models trained on high-quality images generally outperform those trained on low-quality images. It is important to note that when we trained models on high-quality images and tested them on low-quality images, and vice versa, there was an increase in performance. This improvement may be partly due to the larger volume of data available in the training set. We did not need to split the data into training and testing subsets.

#### Limitations

Although we reached out to Google and confirmed that the DDI and ISIC datasets were not used in their training process, the ambiguity surrounding the input data for proprietary, closed-source models still raises concerns about potential data leakage, which could upward bias the predictive performance of Derm Foundation. Moreover, while we have established that skin tone is a key factor in predictive performance, we are constrained by the limited availability of high-quality images with annotated Fitzpatrick Skin Type labels. Consequently, our results are only presented using the DDI dataset. Finally, while our framework is applicable to general medical diagnostic tasks, our initial results have specifically focused on two common modalities in dermatology: dermoscopy images and teledermatology photos. The generalizability of our findings needs to be validated in other medical domains and applications.

#### Future Works

Several promising pathways lie ahead for future research. First, we could develop a comprehensive suite of datasets and models that serve as a “leaderboard” for evaluating medical AI models, initially focusing on dermatological applications. Additionally, armed with the insight that image quality, skin tone, and image-text paired pre-training are likely key differentiators for downstream prediction performance, we plan to enhance foundation models by incorporating multimodal medical data beyond images, such as patient metadata and history, especially for historically underrepresented groups like patients with dark skin tones. Finally, we aim to extend and assess this framework in other medical fields such as cardiology, radiology, and pathology, where there is an abundance of multimodal data and foundation models are beginning to show promise (Huang et al., 2023; Zhang et al., 2023; Chen et al., 2024).

## Data Availability

All code and results reproduced in the present study will be released on github and are available upon reasonable request to the authors before then.

### Appendix A. Details on other vision embedding models

We consider 80 vision models in total:

- 2 domain-specific Dermatology models that are pretrained natural image-text data and then finetuned on medical data: MONET (Kim et al., 2023) and Google Derm Foundation (Steiner, 2024).
- 78 general-purpose vision models:
  - 17 ViT models ranging from ViT-tiny to ViT-giant, trained on supervised and unsupervised tasks including ImageNet-21k (Dosovitskiy et al., 2020) classification, Masked Autoencoders (He et al., 2021), DINO (Caron et al., 2021), and CLIP (Radford et al., 2021), including some finetuned on ImageNet-1k.
  - 1 randomly initialized ResNet-50.
  - 11 ResNet-50 models (He et al., 2016) trained with contrastive learning (Chen et al., 2020) on ImageNet-1k, Places-365 (Zhou et al., 2017; López-Cifuentes et al., 2020), and 9 synthetic image datasets used in Baradad et al. (2022).
  - 49 ResNet-18 models trained with Alignment and Uniformity contrastive loss (Wang and Isola, 2020) on ImageNet-100, Places-365, and 47 realistic and synthetic image datasets from Baradad et al. (2021).

